# Networks and genes modulated by posterior hypothalamic stimulation in patients with aggressive behaviours: Analysis of probabilistic mapping, normative connectomics, and atlas-derived transcriptomics of the largest international multi-centre dataset

**DOI:** 10.1101/2022.10.29.22281666

**Authors:** Flavia Venetucci Gouveia, Jürgen Germann, Gavin JB Elias, Alexandre Boutet, Aaron Loh, Adriana Lucia Lopez Rios, Cristina V Torres Diaz, William Omar Contreras Lopez, Raquel CR Martinez, Erich T Fonoff, Juan C Benedetti-Isaac, Peter Giacobbe, Pablo M Arango Pava, Han Yan, George M Ibrahim, Nir Lipsman, Andres M Lozano, Clement Hamani

## Abstract

Deep brain stimulation targeting the posterior hypothalamus (pHyp-DBS) is being investigated as treatment for refractory aggressive behaviour, but its mechanisms of action remain elusive. We conducted an integrated imaging analysis of a large multi-centre dataset, incorporating volume of activated tissue modeling, probabilistic mapping, normative connectomics, and atlas-derived transcriptomics. 91% of the patients responded positively to treatment, with a more striking improvement recorded in the pediatric population. Probabilistic mapping revealed an optimized surgical target within the posterior-inferior-lateral posterior hypothalamic area and normative connectomic analyses identified fiber tracts and interconnected brain areas associated with sensorimotor function, emotional regulation, and monoamine production. Functional connectivity between the target, periaqueductal gray and the amygdala – together with patient age – was highly predictive of treatment outcome. Finally, transcriptomic analysis showed that genes involved in mechanisms of aggressive behaviour, neuronal communication, plasticity and neuroinflammation may underlie this functional network.

**SIGNIFICANCE STATEMENT:** This study investigated the brain mechanisms associated with symptom improvement following deep brain stimulation of the posterior hypothalamus for severe and refractory aggressive behavior. Conducting an integrated imaging analysis of a large international multi-center dataset of patients treated with hypothalamic deep brain stimulation, we were able to show for the first time that treatment is highly efficacious across various patients with an average improvement greater than 70%. Leveraging this unique dataset allowed us to demonstrate that some patient characteristics are important for treatment success, describe the optimal target zone for maximal benefit, that engagement of distinct fiber tracts and networks within the emotional neurocircuitry are key for positive outcome, and - using imaging transcriptomics - elucidate some potential molecular underpinnings. The provided optimal stimulation site allows for direct clinical application.

## INTRODUCTION

Aggressive behaviours are highly prevalent among psychiatric patients, presenting a major obstacle to patient care. In addition to suffering, these symptoms constitute a leading cause for institutionalization (1–3). Standard treatments for aggressive behaviours involve behavioural and pharmacological therapies that mainly act on the dopaminergic and serotonergic systems (e.g. serotonin reuptake inhibitors, antipsychotics) (1–3). Despite their efficacy, a substantial proportion (30%) of patients fail to respond and are considered to be treatment refractory (4–6). For these patients, neuromodulation therapies, such as deep brain stimulation (DBS), have been investigated as a possible avenue of treatment (2, 7–11). DBS is a neurosurgical therapy in which implanted electrodes are used to adjustably deliver electrical current to specific brain targets (12). It is a well established treatment for movement disorders such as Parkinson’s Disease, dystonia, and essential tremor, and has recently demonstrated promise for several psychiatric and neurological disorders, including Alzheimer’s disease (13, 14), epilepsy (15), obsessive-compulsive disorder (16, 17), depression and anorexia nervosa (18–21).

To date, DBS for refractory aggressive behaviour has primarily targeted the posterior hypothalamus (pHyp-DBS) (2, 7–11) The hypothalamus is a diencephalic structure largely known for its role in controlling homeostasis and motivated behaviours. Along the anterior-posterior axis, the hypothalamus can be divided in three regions (i.e. anterior, medial and posterior) with distinct cell types, projections and functions (2, 22). Preclinical studies have demonstrated that stimulation of the lateral hypothalamus and the ventromedial hypothalamic nucleus, results in altered patterns of aggressive behaviours (23–25). These areas have extensive reciprocal connections with the amygdala and periaqueductal gray, which help modulate the intensity and duration of this behaviour (2, 26). Similar to preclinical findings, DBS and lesions of the posterior hypothalamus have also been shown to reduce aggressive symptoms in humans (for a detailed review see Gouveia et al 2019 (2)). Several international centers have reported on their experience with pHyp-DBS for the treatment of aggressive behaviour with variable outcomes (7–11). Overall, 21 patients diagnosed with autism spectrum disorder, intellectual disability, obsessive-compulsive disorder, epilepsy and schizophrenia with ages ranging from 10-51y have been reported. Using validated scales, improvement in aggressive behaviours at long-term (up to several years of follow-up) was in the order of 38-100% compared to baseline (7–11).

Although case studies and small case series provide valuable insight into the safety and therapeutic impact of pHyp-DBS in individual patients, they do not allow the characterization of clinical phenotypes, optimal stimulation target or insights into brain networks underlying efficacious treatment. In this work, we gathered data from the largest international multi-centre dataset of patients treated with pHyp-DBS for aggressive behaviours to retrospectively investigate possible neurobiological mechanisms of action. Combining a well-documented electrode localization and volume of activated tissue (VAT) modeling pipeline (https://www.lead-dbs.org/) with probabilistic sweet-spot mapping (18, 27), normative connectomics (28–30), and transcriptomics analysis (1, 31) (https://alleninstitute.org/) we delineated a potentially ‘optimized’ surgical target and identified the brain networks and underlying neurobiological processes that might underpin successful pHyp-DBS. Demographic data and pre- and post-operative magnetic resonance imaging (MRI) and computed tomography (CT) scans were obtained from each participant for DBS lead localization, followed by estimation of the VAT and determination of the most efficacious region of stimulation. The VATs were further processed for connectomics analyses to investigate the structural (i.e. fiber tracts) and functional (i.e. brain areas) maps associated with symptom improvement. Using demographics and individual functional connectivity, we tested a predictive model of improvement following pHyp-DBS. Finally, we investigated genes with a spatial pattern of distribution similar to the functional connectivity map to explore associated biological processes. A graphical summary of the methodology used in this study can be found in Figure 1.

**Figure 1.**
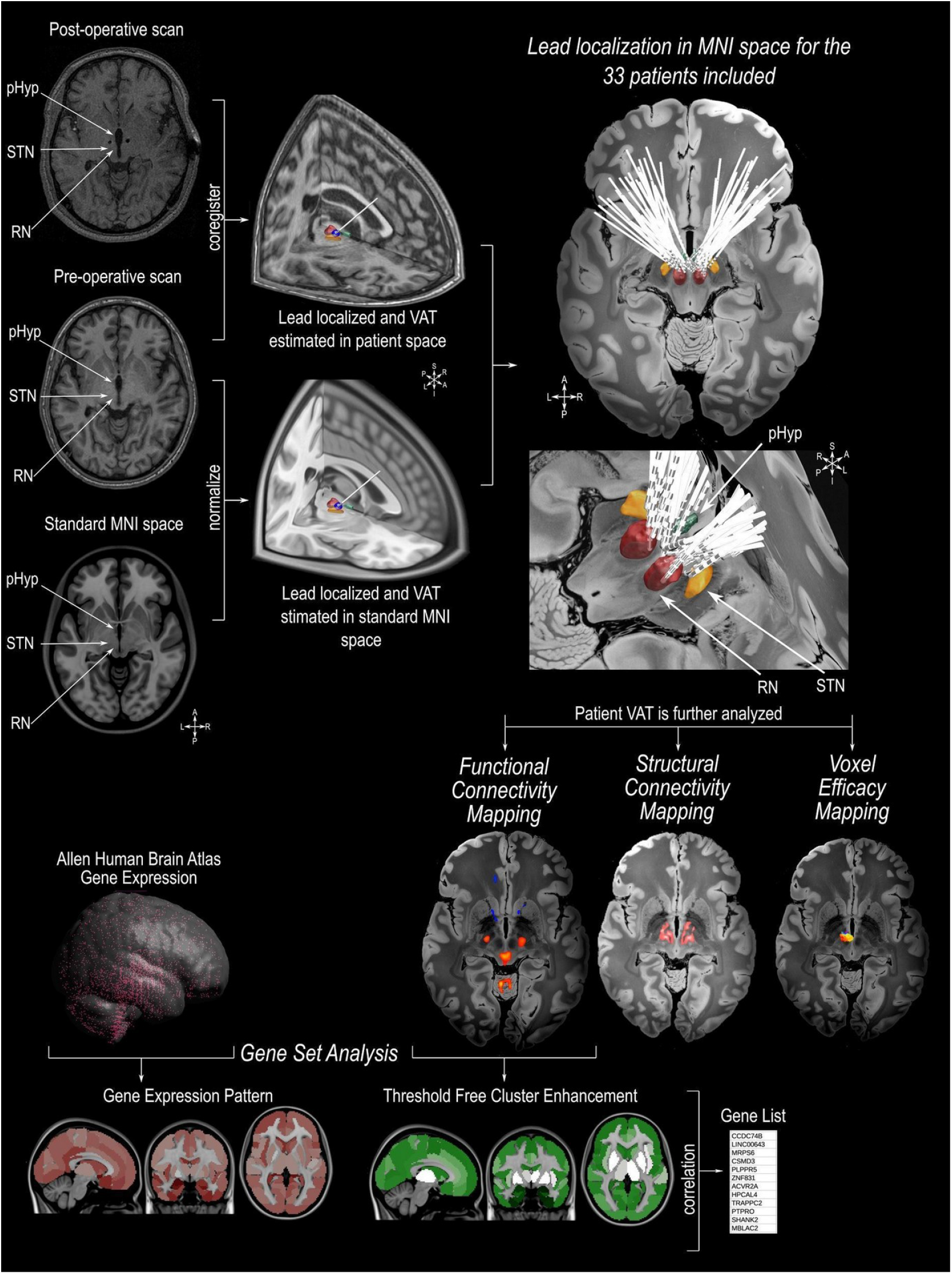
Illustration of the methodologies applied in this study. Preoperative MRI scans were co-registered to the postoperative MRI/CT scan followed by normalization to standard MNI152 space (https://www.bic.mni.mcgill.ca/ServicesAtlases/ICBM152NLin2009). Individual DBS leads were then manually localized in the posterior hypothalamic area (pHyp) in patient space and normalized to MNI152 space. The estimation of the volume of activated tissue (VAT) was calculated based on individual stimulation parameters using Lead-DBS (https://www.lead-dbs.org/ See Table 1 for individual stimulation parameters). The patients’ VATs were further investigated for analysis of the Voxel Efficacy Map (determination of the optimal site of stimulation), Imaging Connectomics using Structural Connectivity Map (determining the fiber tracts involved in symptom improvement) and Functional Connectivity Map (determining the functionally connected areas involved in symptom improvement). For imaging Transcriptomics, we applied a Threshold Free Cluster Enhancement (TFCE) to the functional connectivity map (voxel-wise false-discovery-rate [FDR] correction of q<0.0001). Functional connected areas were averaged into the Harvard-Oxford Atlas (http://www.cma.mgh.harvard.edu/). Based on the human gene expression data from the Allen Human Brain Atlas (https://alleninstitute.org/) genes with a spatial pattern distribution similar to the TFCE map were selected for further gene ontology analysis. 3D reconstruction of the DBS leads on a 100 micron resolution, 7.0 Tesla FLASH brain (https://openneuro.org/datasets/ds002179/versions/1.1.0) in MNI152 space; the pHyp label was derived from a previously published high-resolution MRI atlas of the human hypothalamic region (https://zenodo.org/record/3903588#.YHiE7pNKiF0).

## RESULTS

### Patients Included

In this retrospective study, we aggregated a large dataset of 33 patients from 5 centers treated with pHyp-DBS for alleviation of intractable aggressive behaviours, which were hallmarked by self-injurious behaviour and extreme aggressive behaviour towards patients’ surroundings and others (12 females, 24.48±10.28y of age ranging from 10 to 52y, Figures 2A-B, detailed demographics are found in Table 1). Individual trials and cases were evaluated by the corresponding local ethics committee and informed consent was obtained (7–11, 32, 33). Surgical treatment was approved on a humanitarian basis given the chronicity and severity of symptoms and lack of response to conservative treatment. Whole-brain T1-weighted magnetic resonance imaging (MRI) was acquired preoperatively for surgical planning while a postoperative brain MRI or computed tomography (CT) was obtained for electrode localization. Aggressive behaviour was assessed using standard questionnaires (i.e., Overt Aggressive Behaviour [OAS]; Modified Overt Aggressive Behaviour [MOAS]; Inventory for Client and Agency Planning [ICAP]). Treatment response is reported as the percentage of improvement at last follow-up, in relation to baseline (preoperative) measures. Patients presenting more than 30% improvement were considered to be responders to treatment.

**Table 1.**
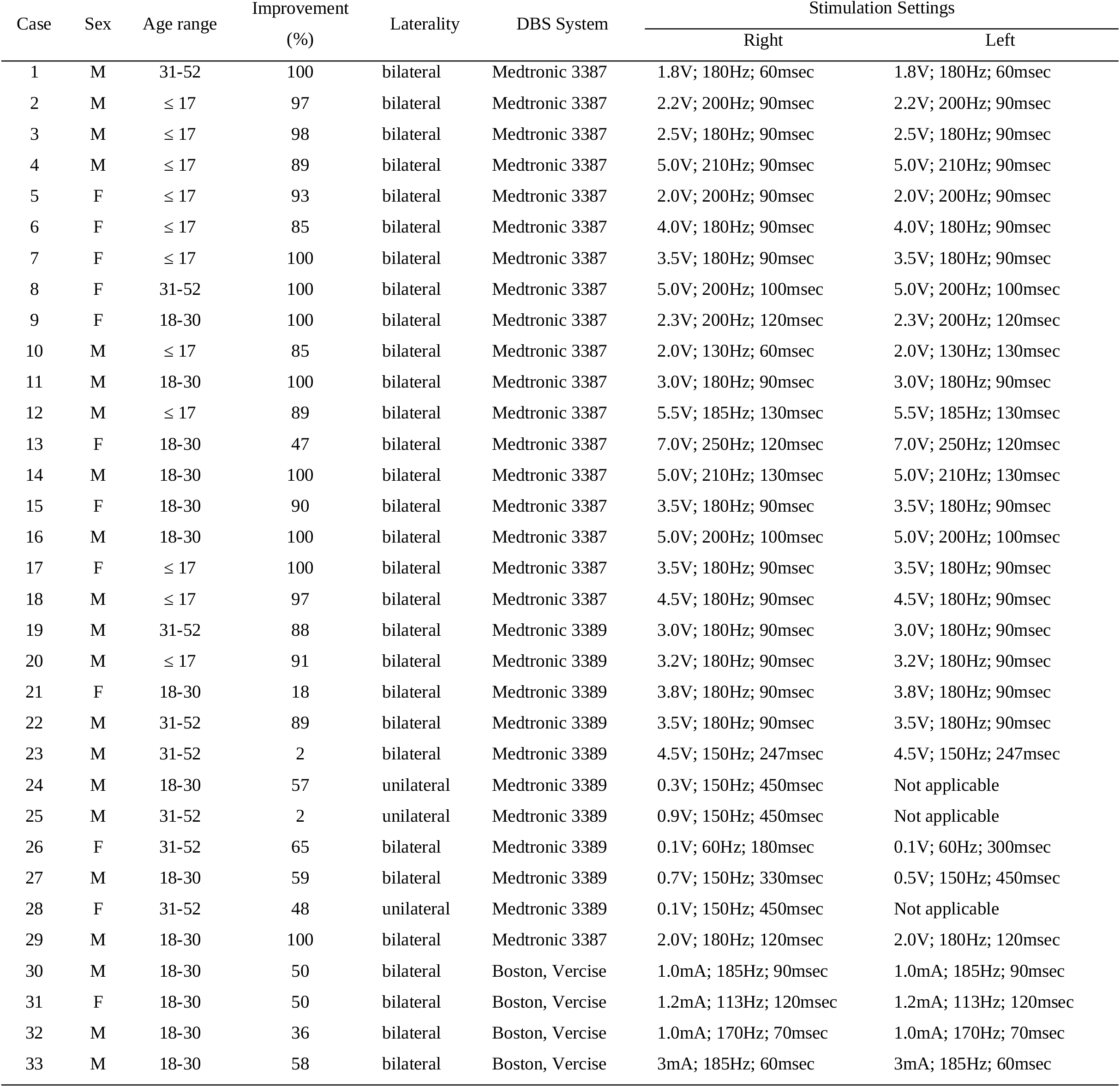
Demographics.

**Figure 2.**
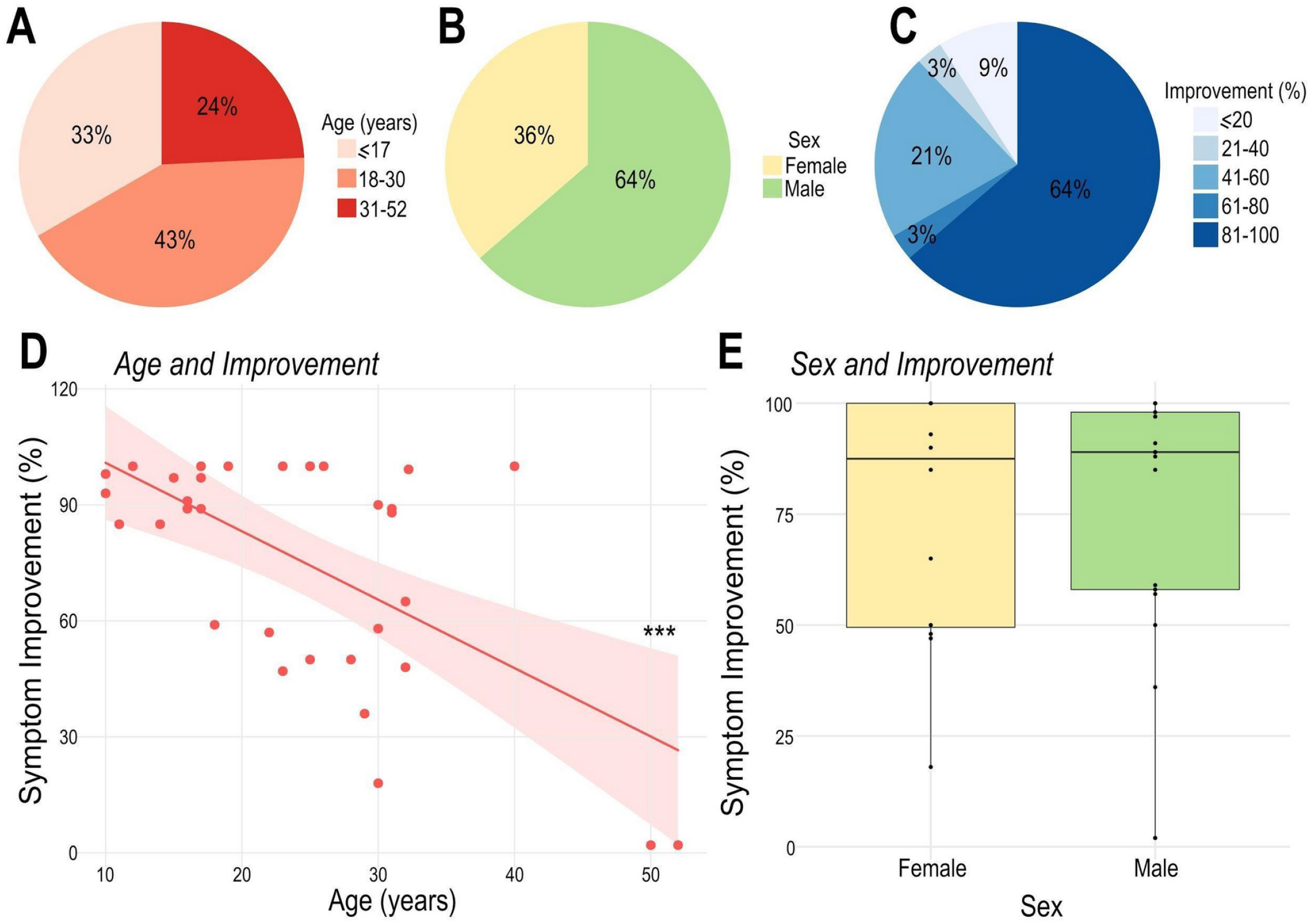
Patient demographics and treatment outcome. A. Patients were divided into three main groups according to age: pediatric population (≤17 years, 11 out of 33), young adults (18 to 30 years, 14 out of 33) and older adults (31 to 52 years, 8 out of 33). B. Distribution of males (21 out of 33) and females (12 out of 33) in this study. C. Patient distribution according to the percentage of symptomatic improvement (⪬20: 3 out of 33; 21-40: 1 out of 33; 41-60: 7 out of 33; 61-80: 1 out of 33; 81-100: 21 out of 33). Note that most individuals had >80% improvement after DBS. D. Age at surgery was significantly negatively correlated with postoperative symptomatic improvement (R= -0.61; R2= 0.38; *** p<0.001). E. There was no significant difference in the percentage of symptom improvement between male and female patients.

pHyp-DBS was implanted bilaterally in 30 patients and unilaterally in 3 using either the Medtronic Activa (3387 leads in 19 cases and 3389 leads in 10 cases) or Boston Vercise DBS Systems (4 cases). Average stimulation parameters for the right hemisphere were 2.45V ± 1.76V (range 0.3 to 4.5V), 166.25Hz ± 18.87Hz (range 150 to 185 Hz) and 219.25μsec ± 172.46μsec (range 60 to 450μsec). For the left hemisphere, average stimulation parameters were 3.17V ± 1.26V (range 2 to 4.5V), 171.67Hz ± 18.93Hz (range 150 to 185 Hz) and 142.33μsec ± 95.48μsec (range 60 to 257μsec). Individual stimulation parameters are shown in Table 1.

After treatment, 91% (30 out of 33) of the patients were considered to be responders (4–6, 9). The average percentage of improvement was 75.25 ± 29.59% (Figure 2C). Younger patients were found to have a more pronounced benefit, with the pediatric population (patients ≤17 years of age) exhibiting greater symptom improvement compared to the adult population (93% vs 66%, Figure 2D). No differences were observed between males (75.57% ± 31.13%) and females (74.68% ± 28.00%; Figure 2E).

### Probabilistic Sweet-spot Mapping

To provide insight into the relationship between stimulation location and response to pHyp-DBS treatment, probabilistic maps of efficacious stimulation were generated using previously described methods (18, 27). Briefly, preoperative MRI scans were co-registered to individual postoperative MRI/CT scans, normalized to standard MNI152 space for estimation of the VAT. Left-sided VATs were flipped at the sagittal plane, weighted by the corresponding percentage of improvement and the mean improvement of overlapping VATs was calculated at each voxel. These average maps were then thresholded for voxel-wise significance using a Wilcoxon signed rank test (p<0.05). Additionally, to exclude outlier voxels, only those included in >10% of all maps were included. Finally, we performed nonparametric permutation testing, randomly assigning each clinical score to a VAT as previously described (27, 34, 35)

This analysis revealed greater symptom alleviation related to stimulation of a more posterior-inferior-lateral region of the posterior hypothalamic area (Figure 3). This area encompassed 684 voxels associated with behavioural improvement greater than 90%. The centroid of this highly efficacious area can be found at x=6.5, y=-16, z=-1.5 in the Talairach-Tournoux space (http://www.talairach.org/), and in MNI152 space at the coordinates x=7.5, y=-15, z=-6.5 (https://www.bic.mni.mcgill.ca/ServicesAtlases/ICBM152NLin2009). The permutation test showed that this pattern indeed reflects the specific relationship of individual VATs with individual outcome (p_permute_<0.01).

**Figure 3.**
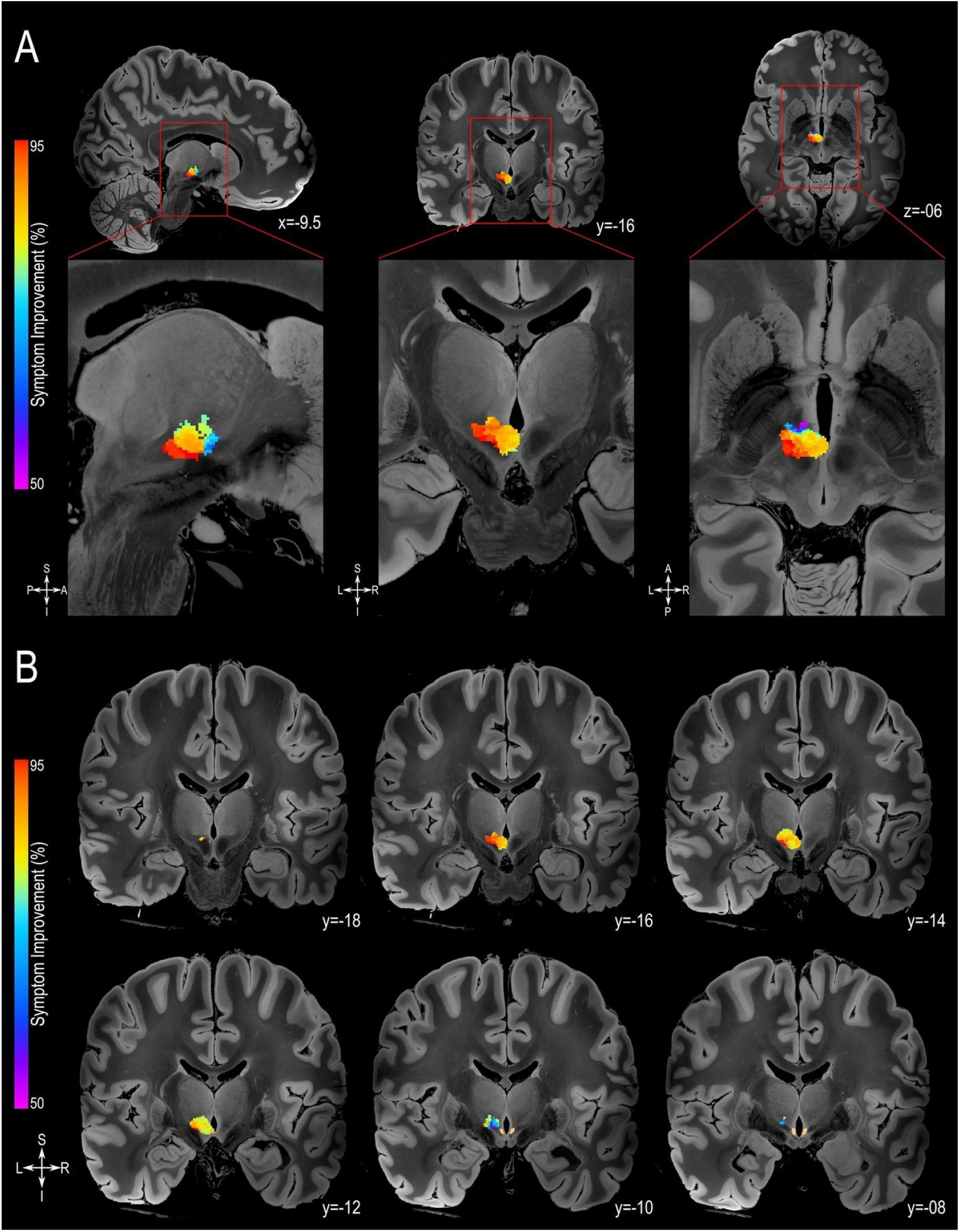
Probabilistic Sweet-spot Mapping. A. The area of stimulation corresponding to greater symptom improvement (red) was located in the more posterior-inferior-lateral region of the posterior hypothalamic area (from left to right: sagittal, coronal and axial views). B. The extent of the volumes of activated tissue (VATs) that were responsible for eliciting at least 50% improvement in symptoms is shown in successive coronal MRI slices. All results are illustrated on slices of a 100 micron resolution, 7.0 Tesla FLASH brain (https://openneuro.org/datasets/ds002179/versions/1.1.0) in MNI152 space (https://www.bic.mni.mcgill.ca/ServicesAtlases/ICBM152NLin2009). The posterior hypothalamic nucleus (pHyp n.) label (shown in beige) was derived from a previously published high-resolution MRI atlas of the human hypothalamic region (https://zenodo.org/record/3903588#.YHiE7pNKiF0).

### Normative Connectomics Analyses - Structural and Functional Connectivity Mapping

To investigate white matter tracts and brain networks associated with symptom improvement, normative structural and functional connectivity mapping were performed ^(28–30)^. Structural connectivity mapping employed diffusion MRI-based tractography data sourced from Human Connectome Project subjects to identify streamlines that intersected individual VATs, while functional connectivity mapping used Genomics Superstruct Project-derived resting-state functional MRI information to generate voxel-wise correlation maps that reflected each VTA’s brain-wide functional connectivity. Subsequent statistical analyses were conducted to determine which streamlines and functional connectivity patterns were associated with positive treatment outcomes.

A variety of streamline bundles were identified by this analysis as being clinically relevant, such that VATs touching said streamlines corresponded to better outcome than VATs that did not. These streamline bundles may be divided into 3 main categories of function: I. Somatosensation (Medial Lemniscus and Spinothalamic Tract); II. Regulation of emotions (Amygdalofugal Pathway, Anterior Limb of the Internal Capsule, Medial Forebrain Bundle (36, 37)); III. Motor Connections (Superior Cerebellar Peduncle, Rubrospinal tract, Frontopontine tract, Central Tegmental Tract, Medial-longitudinal Fasciculus, Motor Projections; Figure 4A-B, Supplementary Figure 1). Interestingly, when overlapping the voxel efficacy and the structural connectivity maps (Figure 4C), we observed that voxels associated with higher efficacy (in red) were more closely related to these fiber tracts.

**Figure 4.**
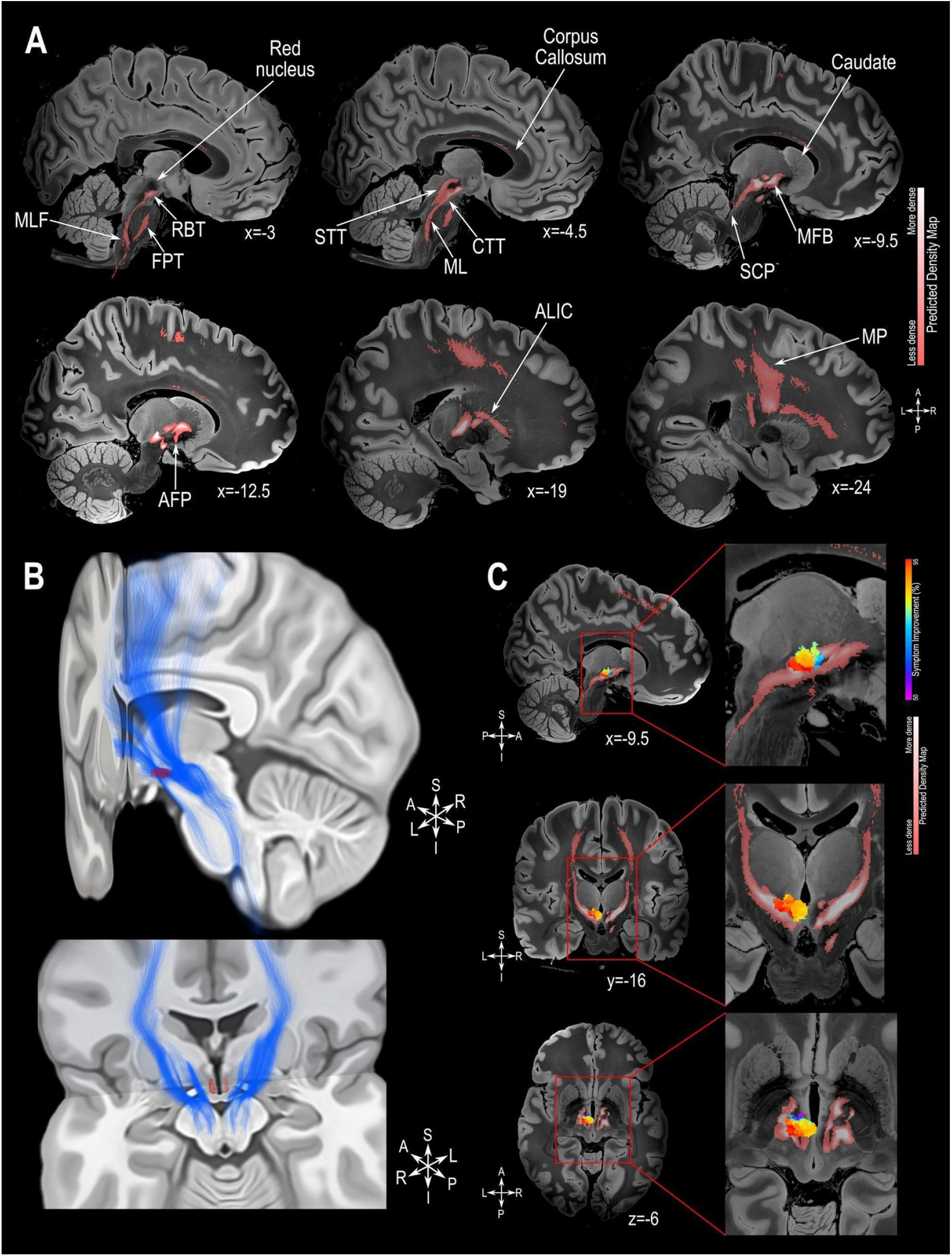
Structural Connectivity Mapping. A. Magnetic Resonance Imaging (MRI) in the sagittal plane showing the fiber density of streamlines significantly connected to the volumes of activated tissue (VATs) associated with greater symptom improvement. B. 3D reconstruction of the streamlines associated with high efficacious VATs illustrated on the MNI152 brain (https://www.bic.mni.mcgill.ca/ServicesAtlases/ICBM152NLin2009); the posterior hypothalamic nucleus label (in red) was derived from a previously published high-resolution MRI atlas of the human hypothalamic region (https://zenodo.org/record/3903588#.YHiE7pNKiF0). C. MRI showing the relation between VATs responsible for eliciting at least 50% improvement in symptoms and the fiber density map (from top to bottom: sagittal, coronal and axial views). The results presented in A and C are illustrated on a 100 micron resolution, 7.0 Tesla FLASH brain (https://openneuro.org/datasets/ds002179/versions/1.1.0) in MNI152 space. Abbreviations: AFP: Amygdalofugal Pathway; ALIC: Anterior Limb of the Internal Capsule; CTT: Central-Tegmental Tract; FPT: Frontopontine Tract; MFB: Medial Forebrain Bundle; ML: Medial Lemniscus; MLF: Medial-Longitudinal Fasciculus; MP: Motor Projections; RBT: Rubrospinal Tract; SCP: Superior Cerebellar Peduncle; STT: Spino-Thalamic Tract.

The functional connectivity analysis showed that the extent of VAT connectedness to several areas was significantly associated with clinical benefits. These areas are related to the production of monoamines (i.e. dorsal and medial raphe nuclei [serotonin]; substantia nigra [dopamine]; Figure 5) and are known to be components within the neurocircuitry of aggressive behaviour (e.g. amygdala; nucleus accumbens, rostral anterior cingulate cortex; bed nucleus of the stria terminalis; hypothalamus; dorsal anterior cingulate cortex, insula, periaqueductal grey; Figure 5) (1, 2, 5, 6). Functional connectivity mapping after Threshold-Free Cluster Enhancement (TFCE) analysis FDR corrected at q<0.0001 is presented in Supplementary Figure 2.

**Figure 5.**
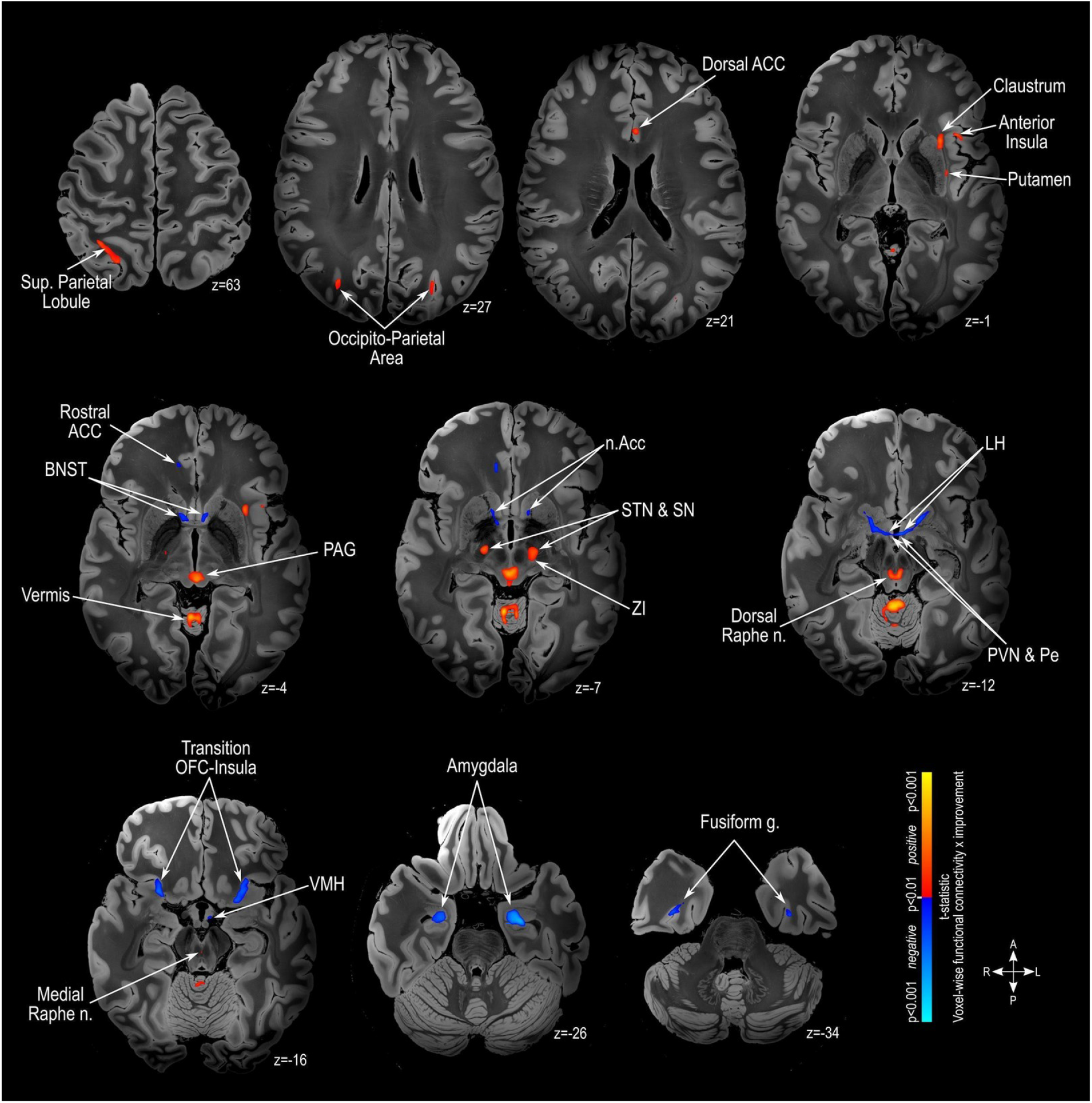
Functional Connectivity Mapping. Magnetic Resonance Imaging (MRI) in the axial plane showing areas found to have positive outcome-connectivity (warm colors) or negative outcome-connectivity (cold colors). The results are illustrated on a 100 micron resolution, 7.0 Tesla FLASH brain in MNI152 space (https://openneuro.org/datasets/ds002179/versions/1.1.0). These results have a significance level of p<0.001 uncorrected. Supplementary Figure 2 shows the results of the functional connectivity mapping after Threshold-Free Cluster Enhancement (TFCE) analysis FDR corrected at q<0.0001. Abbreviations: ACC: Anterior Cingulate Cortex; BNST: Bed Nucleus of Stria Teminalis; LH: Lateral Hypothalamus; n.Acc: Nucleus Accumbens; OFC: Orbitofrontal Cortex; PAG: Periaqueductal Grey; Pe: Periventricular Hypothalamus; PVN: Paraventricular Hypothalamus; SN: Substantia Nigra; STN: Subthalamic Nucleus; VMH: Ventromedial Hypothalamus; ZI: Zona Incerta.

### Estimation of Clinical Outcome

To investigate whether individual functional connectivity to particular hubs within the neurocircuitry of aggressive behaviour could be used to estimate symptom improvement following pHyp-DBS, additive linear models were created. For this, we selected 2 key brain areas that have in previous literature been described as key areas within the neurocircuitry of aggressive behaviour, namely amygdala and periaqueductal gray matter *(1, 2)*. The best performing parsimonious model, which incorporated patient age as well as individual VAT connectivity to the left amygdala and the periaqueductal gray, significantly predicted approximately half of the variance in individual symptom improvement (R=0.7, R^2^=0.49, p<0.00001) and retained significance during leave one-out cross-validation (LOOCV) (R=0.59, R^2^=0.35, p=0.00029; Figure 6).

**Figure 6.**
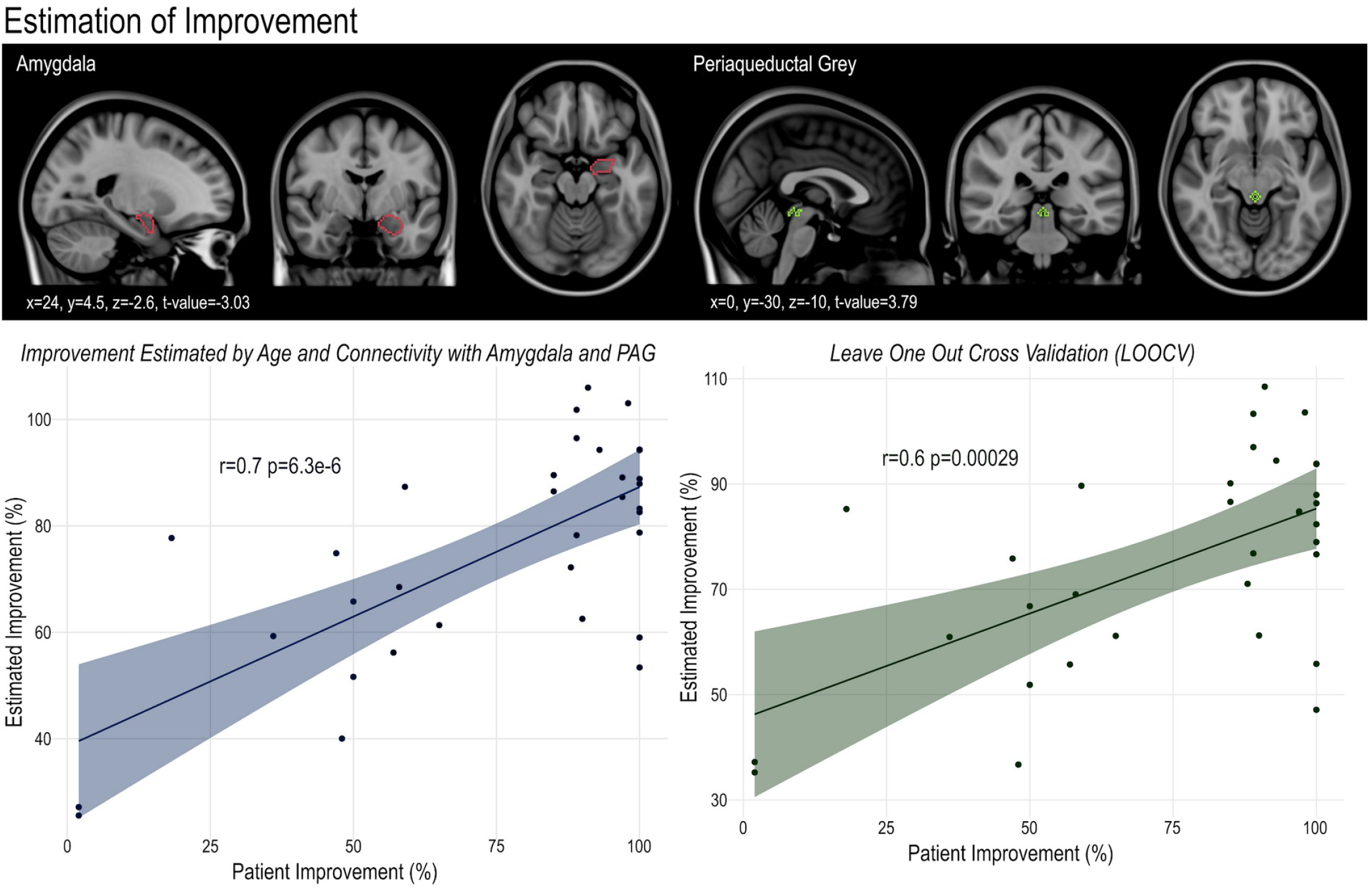
Estimation of clinical outcome. A. Age and individual VAT connectivity to the left amygdala and to the periaqueductal gray significantly estimated individual symptom improvement in the whole dataset (R=0.7, R^2^=0.49, p<0.00001; I) and in leave one-out cross-validation (LOOCV) approach (R=0.59, R^2^=0.35, p=0.00029; II). Bottom panels show the region of interest for the left amygdala (red outline) derived from the Harvard-Oxford Atlas (www.cma.mgh.harvard.edu), and the region of interest for the periaqueductal gray (green outline) derived from the Harvard Ascending-Arousal Network Atlas (https://www.nmr.mgh.harvard.edu/resources/aan-atlas), in sagittal, coronal and axial slices (from left to right) and illustrated in MNI152 standard-space (http://www.bic.mni.mcgill.ca/ServicesAtlases/HomePage).

### Imaging Transcriptomics – Gene Set Analysis

Finally, to investigate neural phenotypes and possible neurobiological mechanisms of treatments we performed imaging transcriptomics analysis using human gene expression data from the Allen Human Brain Atlas (https://alleninstitute.org/), we investigated genes with a spatial distribution of expression that resembled the pattern of brain regions with clinically relevant (following TFCE correction) functional connectivity to the stimulation locus (q_FDRcor_<0.0001). This type of analysis shows genes whose spatial pattern correlates with brain changes and are also found in post-mortem and large-scale GWAS studies of specific patient populations (38). This process resulted in an extensive list of candidate genes that were Bonferroni corrected (p<0.005) before being investigated using the EnRichr Gene Ontology tool (https://maayanlab.cloud/Enrichr/) and a cell-specific aggregate gene set (39) to identify associated biological processes and cell types (Figure 7). We observed genes associated with the clinically relevant brain areas considered to be at the core of the neurocircuitry of aggressive behaviour (i.e. Hypothalamus, Amygdala, Prefrontal Cortex and Cingulate Cortex)(1, 2, 5, 6, 40), genes linked to oxytocin (a peptide hormone critically implicated in social behaviours) (41–45) and to cell compartments that are related to neuronal communication and plasticity (e.g. neuron projection, synapse; axon guidance, long-term potentiation). Additionally, investigating the prevalence of cell-types associated with the set of genes identified in our analysis, we found a significant overrepresentation of genes associated with oligodendrocytes and an underrepresentation of genes associated with astrocytes and microglia when compared to the expected distribution of genes per cell type (Figure 7).

**Figure 7.**
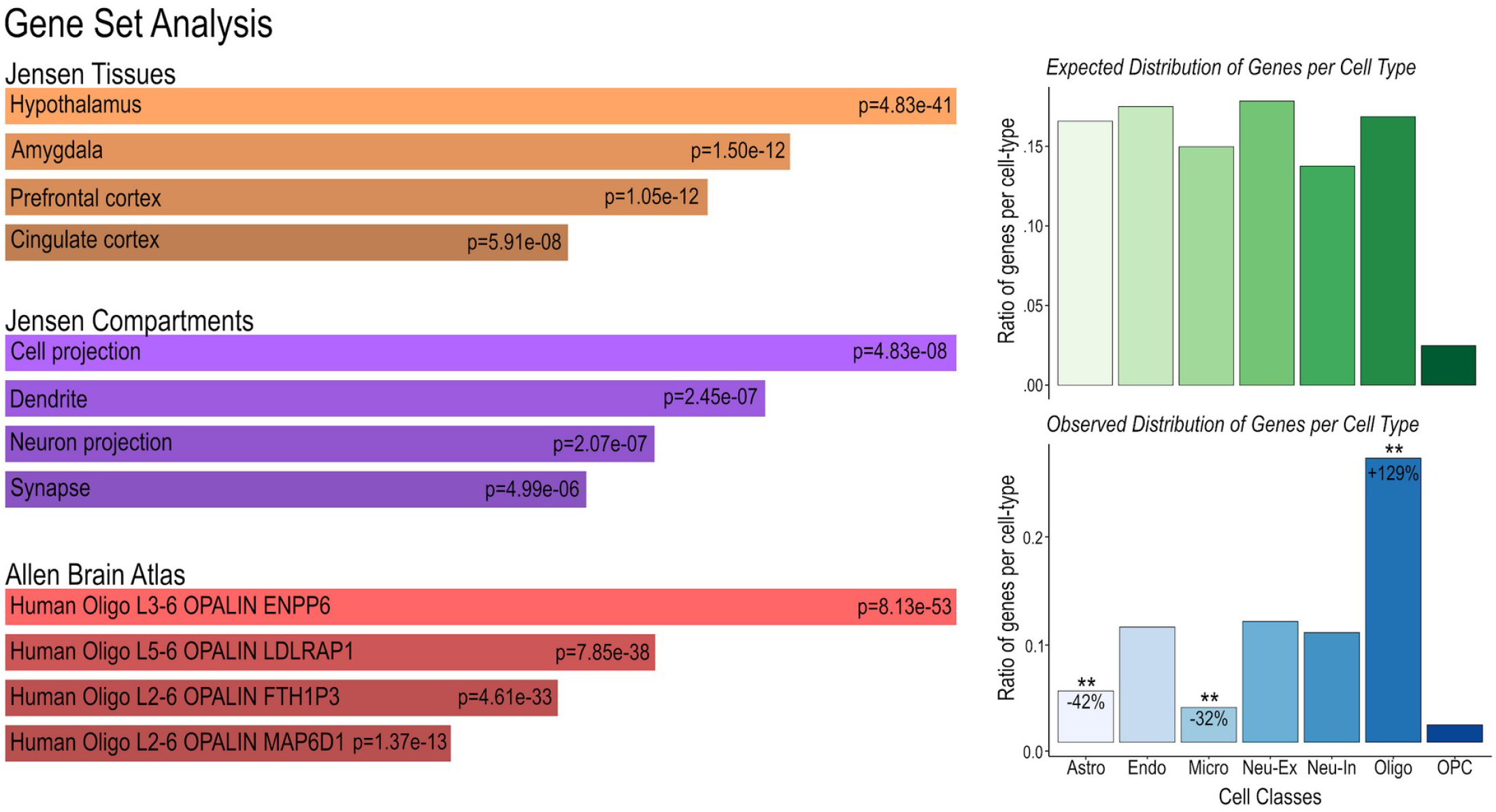
Imaging transcriptomics-gene set analysis. The gene set analysis was performed using the TFCE-corrected distribution of clinically relevant functionally connected areas (q_FDR_<0.0001) along with whole brain three-dimensional expression patterns provided by the Allen Brain atlas (http://human.brain-map.org/)(83–85), both averaged into the Harvard-Oxford Atlas (http://www.cma.mgh.harvard.edu/). Genes with similar spatial pattern distribution to the functional connectivity map were Bonferroni corrected at p<0.005 and selected for further gene ontology analysis. I) The EnRichr tool (https://maayanlab.cloud/Enrichr/) ^(86)^ was used to investigate associated biological processes, followed by specific tissue and compartment analysis provided by the Jensen Gene Ontology Tool (https://jensenlab.org/resources/proteomics/) and the Kyoto Encyclopedia of Genes and Genomes (KEEG; https://www.genome.jp/kegg/). II) A cell-specific aggregate gene set provided by Seidlitz et. al., 2020 (39) was used to determine the cell-types associated with these genes. The results were confirmed to be non-random using permutation testing (1000 permutations, ** p<0.01).

## DISCUSSION

In this work, we performed an integrated imaging analysis of a large, international multi-centre dataset of patients treated with pHyp-DBS for severe and refractory aggressive behaviours. Overall, the response rate for pHyp-DBS was high, with 91% (30 out of 33) of patients qualifying as treatment responders (i.e. 30% improvement in behaviour). In addition, 64% of patients presented symptom improvement greater than 81% (Figure 2). The mean improvement across the whole pHyp-DBS cohort was 75%. These results are in line with previous work showing that symptom improvement following hypothalamic neurosurgery for the treatment of aggressive behaviour is in the order of 90% (2). Interestingly, four of these patients were implanted with directional DBS leads and presented similar symptom alleviation on follow-up as the other patients included here (8). We observed that pediatric patients presented the highest percentage of symptom improvement.

Considering that the brain only fully achieves maturity in early adulthood (46), it is possible that treating dysfunctional circuitries while they are still being developed could induce beneficial and long-lasting plastic changes in the direction of restoring functional normality. DBS is primarily used for the treatment of neurological and psychiatric disorders in adults and is emerging as an effective and safe therapy for neurological diseases in children (47) such as childhood dystonia (48), drug-resistant epilepsy (49) and Gilles de la Tourette syndrome (50). Although analysis derived from retrospective clinical data is more prone to potential bias, this study included a collection of previously published prospective studies, where each participant center standardized the timeframe for data collection and the instruments used to evaluate symptom improvement.

Using patients’ pre and postoperative imaging, we performed a Voxel Efficacy Mapping investigation and were able to determine a specific area that, when stimulated, is associated with improvement greater than 90% located in the more posterior-inferior-lateral aspect of the posterior hypothalamic area (coordinates are provided in Talairach-Tournoux space and MNI152 space in the Results section, Figure 3). In humans, DBS and lesions of the posterior hypothalamus have been shown to reduce aggressive symptoms (for a detailed review see Gouveia et al 2019 (2)). One of the most commonly targeted areas lies along the midpoint of the line between the anterior and posterior commissures (AC-PC line), in a region anterior to the rostral end of the aqueduct, posterior to the anterior border of the mammillary body and superior to the red nucleus (2, 51).

Our connectomics analyses provided interesting insights into the structural and functional networks that underlie the clinical efficacy of pHyp-DBS. As no native functional or diffusion weighted imaging scans were available in the patient population, these analyses were performed using high resolution, high signal-to-noise ratio images derived from healthy individuals (7, 13, 18, 30, 52). Although normative connectivity data may not fully capture patient- or pathology-specific variations, they have been previously shown to generate comparable results to patient-specific imaging data (52). The current study’s structural connectivity analysis implicated fiber tracts involved in somatosensation (medial lemniscus and spinothalamic tract), emotional regulation (e.g., amygdalofugal pathway and medial forebrain bundle), and motor signaling (e.g., motor projections and central tegmental tract) in the amelioration of aggressive behaviour. Indeed, patients with neurodevelopmental disorders, especially those presenting with self-injurious behaviour, are known to often have altered sensory perception (including reduced pain sensitivity), which can contribute to the chronicity and severity of the behaviour (1, 53). As such, modulation of somatosensory pathways – perhaps leading to increased pain perception or awareness – could help to reduce self-injurious behaviours. More generally, aggressive behaviours and motor agitation are believed to occur as a result of a decreased tolerance to provocative stimuli caused by reduced serotonergic transmission in the prefrontal cortex, resulting in an ineffective top-down inhibitory control over a consequently hyper-activated amygdala. The amygdala in turn sends alert signs to the periaqueductal gray and hypothalamus for motor activation and hormonal production, preparing for fight-or-flight responses (1, 2, 5, 6, 40). Thus, it is conceivable that stimulation of fiber tracts involved in emotional regulation and motor outputs could mediate clinical improvement by reducing motor agitation, improving concentration and learning, and increasing patients’ tolerance to provocative external stimuli. Furthermore, the involvement of the main tracts responsible for somatosensation (i.e. medial lemniscus and spinothalamic tract) could explain the reduction in self-injurious behaviours by an increased pain perception.

Functional connectivity mapping showed that the most successful VATs were functionally connected to several areas within the wider neurocircuitry of aggressive behaviour (e.g., amygdala, nucleus accumbens, hypothalamus, periaqueductal gray, and cingulate cortex) as well as areas responsible for the production of serotonin (i.e., raphe nuclei) and dopamine (i.e., substantia nigra). Both these monoamines are thought to be directly involved in the initiation and maintenance of aggressive behaviours due to their role in facilitating top-down inhibitory controls, regulating mood, and mediating social behaviours (2, 7, 54). In fact, antipsychotics (both typical and atypical), and selective serotonin reuptake inhibitors – the most commonly used drugs for the control of aggressive behaviours – act on the dopaminergic and serotonergic neurotransmission systems (1, 2). Confirming the major role of the amygdala and the periaqueductal gray in the neurocircuitry of aggressive behaviour, we showed that VAT connectivity to these areas is a key predictor of individual outcomes following pHyp-DBS. Previous studies have shown positive outcomes following ablative neurosurgery targeting the amygdala for the control of aggressive behaviours (1, 2, 6). DBS of the periaqueductal grey – primarily performed to treat neuropathic pain (55) – has also been reported to modulate mood and anxiety (56). Altogether, the connectomics analysis performed here expands our understanding on the neurocircuitry of aggressive behaviour, and suggests that other brain areas within the neurocircuitry are key for symptom alleviation. It would be interesting to investigate if neuromodulation of these deep brain structures with other non-invasive neuromodulatory treatments, such as deep transcranial magnetic stimulation (dTMS), would result in beneficial outcomes for patients.

Imaging transcriptomics is a powerful non-invasive method with which to measure neural phenotypes and investigate neurobiological mechanisms of disease or treatments (38). Genes whose spatial pattern correlates with brain changes in imaging transcriptomics studies are also found in post-mortem and large-scale GWAS studies of specific patient populations (38). In fact, several studies have demonstrated associations between alterations of the spatial distribution of genes and neurodevelopmental (57, 58) and psychiatric disorders (59). In this study, we identified several genes with similar spatial distribution to brain regions that showed clinically important functional connectivity to the stimulation target. These genes can be divided into three groups based on associated biological processes: I) Brain areas or hormones associated with aggressive behaviour; II) Neuronal communication and plasticity; III) Neuroinflammation. Genes whose biological processes are associated with aggressive behaviour include those linked to the limbic system (e.g. amygdala and hypothalamus) and higher-order cognitive centers (e.g. prefrontal cortex). As described above, a dysregulation in the top-down inhibitory control of the prefrontal cortex over the amygdala could lead to aggressive behaviour.

Additionally, we identified genes implicated in the oxytocin signaling pathway, a peptide hormone critically implicated in aggressive behaviour (43). It has been shown that intranasal administration of oxytocin increases human aggressive responses when subjects are evaluated in a social orientation paradigm (41). Moreover, single nucleotide polymorphisms in the oxytocin gene have been associated with childhood-onset aggressive behaviours (44) and with aggressive behaviours in men intoxicated with alcohol (42).

The second group of genes are associated with neuronal communication and plasticity and, thus, are possibly linked to the intrinsic mechanism of action of DBS (60, 61). As a neuromodulation therapy, DBS induces long-lasting changes in cellular and molecular aspects of neurons belonging to dysfunctional neurocircuitries in order to restore functional normality (12, 62, 63). Although its neurobiological mechanisms are not fully understood, DBS is believed to exert its effects by altering the cellular membrane potential resulting in either increased or decreased in action-potentials (12, 62, 63). These changes in local field potential are then propagated throughout the neural network, changing neurotransmitter dynamics, protein expression, and membrane receptor availability (12, 62, 63). In fact, a previous study using imaging transcriptomics to investigate brain changes after functional neurosurgery of the amygdala in patients with refractory aggressive behaviour also identified several genes related to neuronal communication and plasticity (1). Preclinical studies have also found significant increases in neural precursor cells, plasticity, and precursor cell markers in animals implanted with DBS, along with significant reductions in the number of activated microglia and astrocytes (64– 68). This is in line with the cell type investigation that demonstrated a significant reduction in genes associated with astrocytes and microglia when compared to the expected distribution of genes per cell type. Both astrocytes and microglia are highly implicated in mechanisms of neuroinflammation (69), which in turn is associated with several psychiatric disorders (70). The reduction of neuroinflammatory markers (such as glial activation and interleukin levels), has been described in several preclinical DBS studies and is thought to be a key mechanism of action of DBS (66, 71, 72).

## METHODS

### Patients Included

33 patients from five international centers were included in this study: Hospital Universitario San Vicente Fundación, Colombia; University Hospital La Princesa, Spain; International Misericordia Clinic, Colombia; Universidad Autónoma de Bucaramanga and FOSCAL Clinic, Colombia; Sírio-Libanês Hospital, Brazil. Detailed demographics, medical history, surgical procedure, and relevant information regarding institutional ethical review board and informed consent can be found in previous publications (7–11). All patients had pre-operative T1w MRI images as well as postoperative imaging (CT or MRI) that allowed for the precise determination of individual electrode placement (imaging parameters may be found in the original publications of the individual centers) (7–11).

### Electrode localization and Volume of Activated Tissue (VAT) modeling

Lead-DBS software (https://www.lead-dbs.org/) was used for electrode localization and modeling of the volume of activated tissue (VAT), as previously described (73). Following intensity inhomogeneity correction, the postoperative MRI or CT and the preoperative MRI scans were rigidly co-registered using SPM12 (https://www.fil.ion.ucl.ac.uk/spm/software/spm12/ (74)). Images were non-linearly normalized to standard space (ICBM 2009b NLIN asymmetric) using Effective Low Variance ANTS (http://stnava.github.io/ANTs) and an additional subcortical affine transformation was implemented to correct for post-operative brain shift (75). Electrodes located in the right and/or left hemispheres were manually localized using the post-operative scan and then warped to standard space using the previously generated transforms. The patient’s unilateral or bilateral VATs associated with individual stimulation parameters at last follow-up (individual stimulation parameters are described in Table 1) were modeled by first constructing a four-compartment volume conductor model (http://iso2mesh.sourceforge.net/cgi-bin/index.cgi) that segregated peri-electrode tissue by tissue type (76). Next, the FieldTrip-SimBio finite element model pipeline was used to simulate the potential electric field distribution around each active contact (https://www.mrt.uni-jena.de/simbio/index.php/; http://fieldtriptoolbox.org) and binary VATs were generated by thresholding the gradient of this distribution at 0.2 V/mm.

### Voxel efficacy Mapping

Probabilistic maps of efficacious voxels were generated to provide insight into spatial patterns of response to pHyp-DBS, as previously described (18, 27, 30). All VATs were flipped to the right hemisphere and each VAT was weighted by percentage of improvement from baseline. Mean improvement at each voxel was computed by averaging the normalized weighting values. The resultant raw average map was then masked by a frequency map thresholded at 10% in order to exclude outlier voxels. Finally, a statistical map, thresholded at p<0.05, was calculated using Wilcoxon signed-rank tests to determine whether pHyp-DBS was associated with significant difference in clinical change at each voxel. The validity of the voxel efficacy map was confirmed using a non-parametric permutation analysis, in which each clinical score was randomly assigned to a random VAT. The voxel map was determined to be significant (p_permute_<0.01). The coordinates relative to the most efficacious area of stimulation were extracted in MNI space and converted to Talairach space using the MNI ←→ Talairach Converter from BioImage Suite Web (https://bioimagesuiteweb.github.io/webapp/mni2tal.html) (77)

### Imaging Connectomics Analyses - Structural and Functional Connectivity Mapping

The brain-wide imaging connectomics analyses of the patient’s unilateral or bilateral VATs was explored using high-quality Structural (diffusion MRI-based tractography derived streamlines) and Functional (resting-state functional MRI-derived voxel-wise functional pattern) normative connectomes as previously described (18, 29, 30, 78). Briefly, for each patient’s individual VAT(s), a whole-brain r-map was computed based on the resting-state functional MRI BOLD time course-dependent correlations between the seed region and the remaining voxels in the brain (in-house MATLAB script, The MathWorks, Inc., Version R2017b. Natick, MA, USA, the processed rsfMRI connectome data and script are also freely available through lead-dbs - https://www.lead-dbs.org/) across 1000 healthy subjects (Brain Genomics Superstruct Project dataset, age range: 18-35 years; 57% female, http://neuroinformatics.harvard.edu/gsp) (79). To exclude voxels with potentially spurious correlations, all individual r-map were corrected for multiple comparisons by converting them to a t-map and thresholded at p_corrected_<0.05 (whole-brain voxel-wise Bonferroni correction) using the known p-distribution (80, 81). Thus only voxels that are significantly functionally connected to each patient’s VTA are used for the subsequent analysis. Voxel-wise linear regression analysis investigating the relationship between functional connectivity and outcome were then performed. Structural connectivity outputs were obtained by identifying all streamlines that touched the uni or bilateral VATs (used as seeds) out of an approximately 12 million-fibres whole-brain tractography template (in-house MATLAB script, The MathWorks, Inc., Version R2017b. Natick, MA, USA; the processed dMRI connectome data and script are also freely available through lead-dbs -https://www.lead-dbs.org/). This template was assembled from a 985-subject multi-shell diffusion-weighted MRI Human Connectome Project dataset (http://www.humanconnectomeproject.org) using generalized q-sampling imaging (http://dsi-41studio.labsolver.org/). To determine what streamlines are significantly associated with outcome a t-test of symptom improvement comparing subjects whose VAT touched a given streamline and individuals who did not was performed as previously described (17, 30).

To determine the consistency of the results of both structural and functional connectivity mapping, the data was divided in four random groups of even size and the analyses repeated four times leaving out one group each time. The outcome of these four-fold consistency analyses are illustrated in Supplementary Figure 3. While statistical power is reduced due to the smaller sample size in each sub-analysis, the outcomes are highly similar highlighting the same regions and tracts demonstrating that the results reported are consistent across patients.

### Imaging Transcriptomics - Gene Set Analysis

We performed a gene set analysis to investigate genes whose spatial pattern distribution is similar to the pattern of clinically relevant functional connectivity. To this end, we applied Threshold Free Cluster Enhancement (TFCE) to the functional connectivity map (voxel-wise false-discovery-rate [FDR] correction of q<0.0001) and functionally connected areas were averaged into the Harvard-Oxford Atlas (http://www.cma.mgh.harvard.edu/). Based on the human gene expression data from the Allen Human Brain Atlas (https://alleninstitute.org/), genes with similar patterns of spatial distribution to the TFCE map were compiled in an extensive list of candidate genes that were Bonferroni corrected (p<0.005) before being investigated using the EnRichr Gene Ontology tool (https://maayanlab.cloud/Enrichr/), along with the Jensen Gene Ontology enrichment tool (https://jensenlab.org/resources/proteomics/), the Kyoto Encyclopedia of Genes and Genomes (KEGG pathway enrichment; https://www.genome.jp/kegg/kegg1b.html) and a cell-specific aggregate gene set (39) to determine associated biological processes and cell types.

### Statistical Analysis

R (version 3.4.4; https://www.r-project.org/) was used for statistical analysis. The RMINC package (https://github.com/Mouse-Imaging-Centre/RMINC) was used for analysis of imaging data. Linear models were used to investigate the percentage of improvement by age and t-tests were performed to investigate possible differences in symptom improvement comparing male and female patients. An additive linear model was used to test our ability to estimate percentage of improvement from baseline using age and the functional connectivity with the left amygdala and periaqueductal grey - key areas within the neurocircuitry of aggressive behaviour (2, 40, 82), followed by leave-one-out cross-validation.

### Study approval

Individual trials and cases were evaluated by the corresponding local ethics committee and written informed consent was obtained. Five international centers shared clinical data for this study.

1. Comité de Ética de la Investigación of Hospital Universitario San Vicente Fundación gave ethical approval for this work.
2. Comité de Ética de la Investigación con Medicamentos of Hospital Universitario La Princesa gave ethical approval for this work.
3. Comité de Ética de los Estudios Clínicos of La Misericordia Clínica Internacional gave ethical approval for this work.
4. Comité Institucional de Ética of Universidad Autónoma de Bucaramanga gave ethical approval for this work.
5. Comitê de Ética em Pesquisa of Sociedade Beneficente de Senhoras Hospital Sírio-Libanês gave ethical approval for this work.

## Data Availability

All data produced in the present study are available upon reasonable request to the authors

## AUTHOR CONTRIBUTIONS

Design of the study: FVG, JG, CH. Acquisition of data: FVG, ALLR, CVTD, RCRM, ETF, JCBI, WOCL. Analysis of data: FVG, JG, GJBE. Interpretation of data: FVG, JG. Writing – original draft: FVG, JG, CH. Writing – review & editing: FVG, JG, GJBE, AB, AL, ALLR, CVTD, WOCL, RCRM, ETF, JCBI, PG, PMAP, HY, GMI, NL, AML, CH. All authors approved the version to be published. FVG and JG contributed equally to this paper and share first authorship.

## ACKNOWLEDGMENTS

This work was supported by funds from the Harquail Center for Neuromodulation, Canadian Institutes of Health Research (CIHR) Postdoctoral Fellowship #472484 (FVG), Fundacao de Amparo a Pesquisa do Estado de Sao Paulo (FAPESP) #13/20602-5 (FVG), Fundacao de Amparo a Pesquisa do Estado de Sao Paulo (FAPESP) #17/10466-8 (FVG), Canadian Institutes of Health Research (CIHR) Banting fellowship #471913 (JG), and Fundacao de Amparo a Pesquisa do Estado de Sao Paulo (FAPESP) #11/08575-7 (RCRM). The authors wish to thank the research assistants and staff of Hospital Sirio-Libanes, Brazil; Hospital Universitario San Vicente Fundación, Colombia; University Hospital La Princesa, Spain; International Misericordia Clinic, Colombia; Universidad Autónoma de Bucaramanga and FOSCAL Clinic, Colombia.

---

**Supplementary Figure 1.**
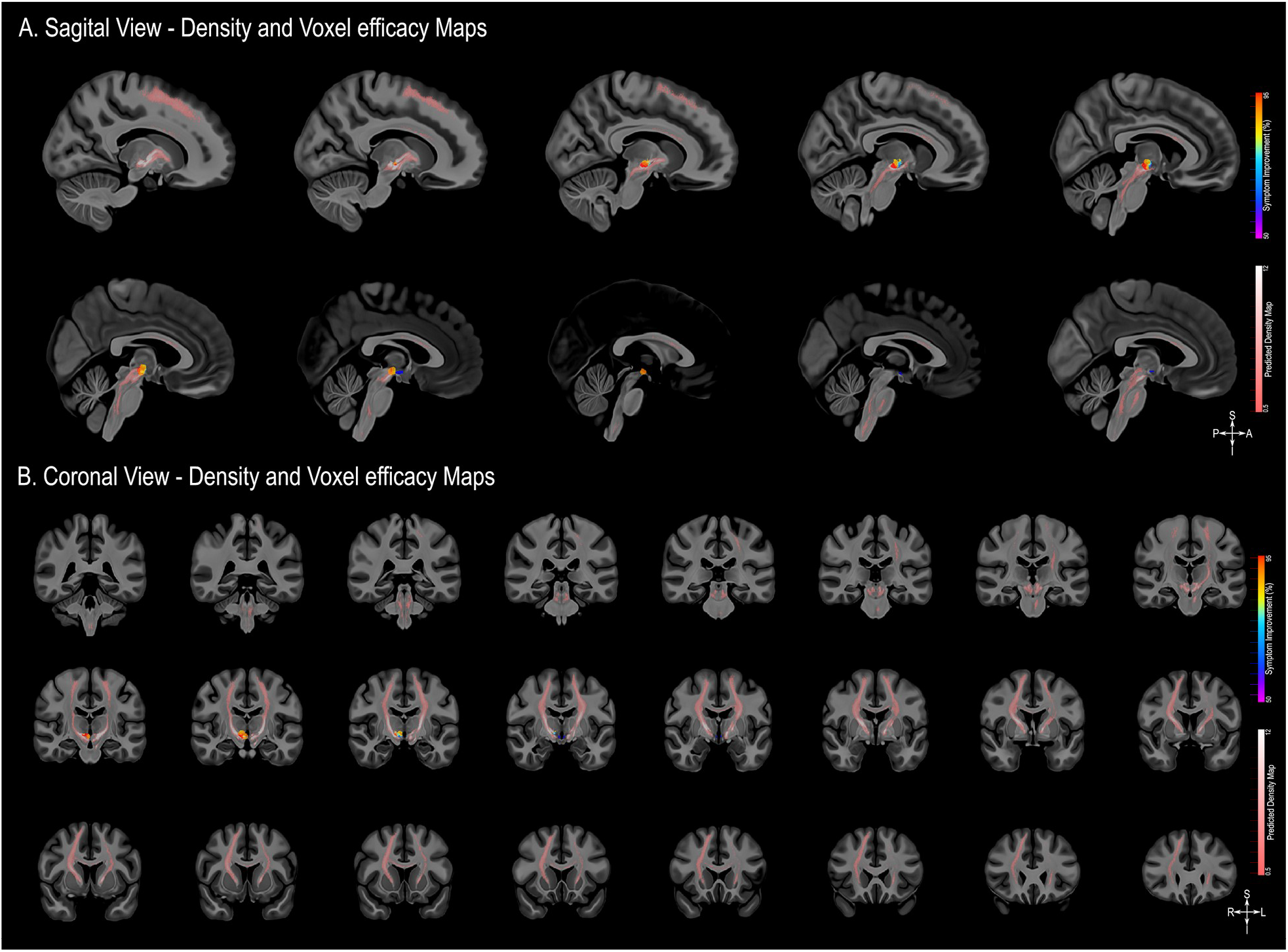
Structural Connectivity Mapping. Magnetic Resonance Imaging (MRI) in the sagittal plane (A) and coronal plane (B) showing the fiber density of streamlines (pink scale) significantly connected to the volumes of activated tissue responsible for eliciting at least 50% improvement in symptoms (heat map) illustrated a 100 micron resolution, 7.0 Tesla FLASH brain (https://openneuro.org/datasets/ds002179/versions/1.1.0) in MNI152 space.

**Supplementary Figure 2.**
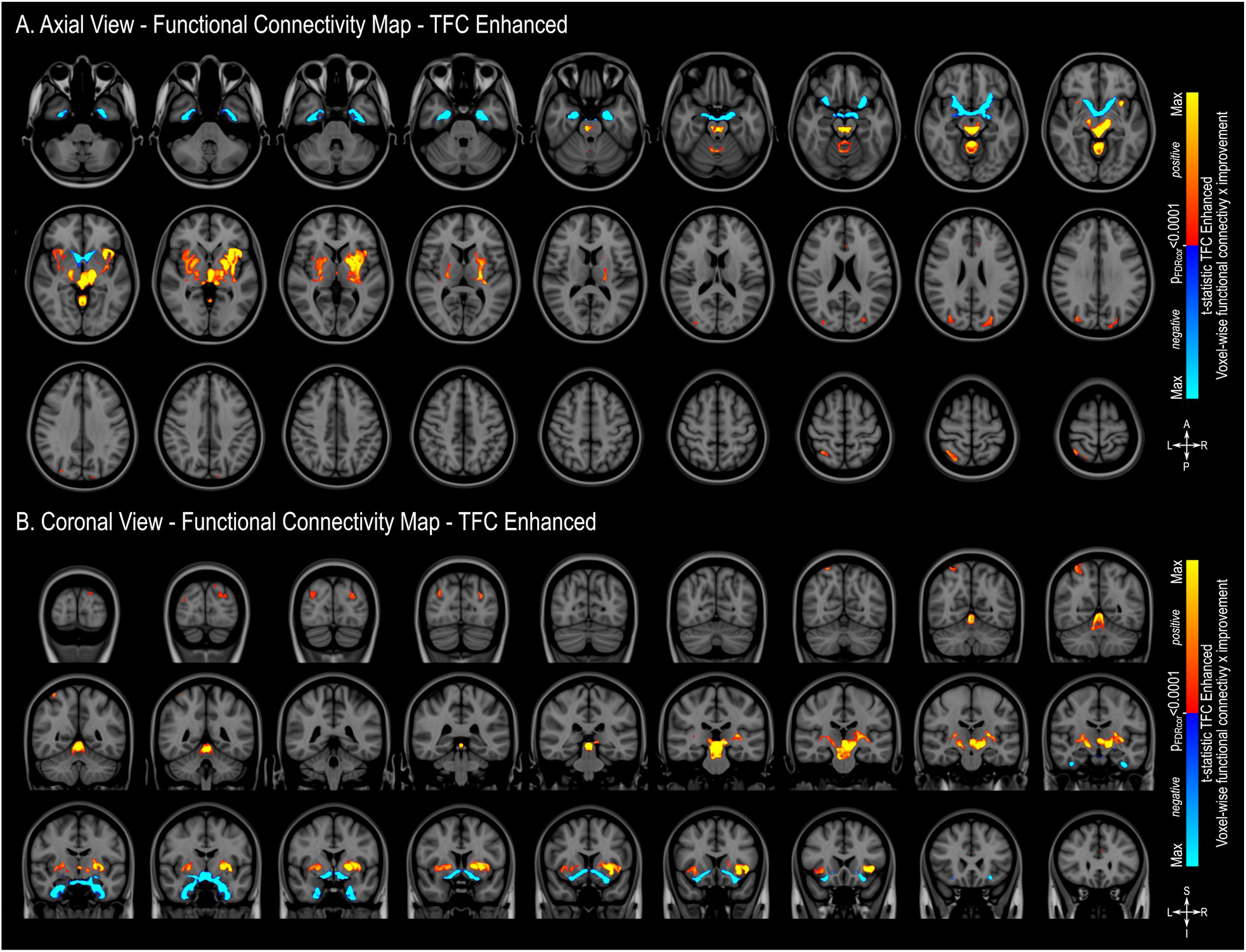
Threshold-free Cluster Enhancement Functional Connectivity Mapping. Magnetic Resonance Imaging (MRI) in the axial plane (A) and coronal plane (B) showing areas found to have positive outcome-connectivity (warm colours) or negative outcome-connectivity (cold colours) FDR corrected at q<0.0001. The results are illustrated in the MNI152 brain (https://www.bic.mni.mcgill.ca/ServicesAtlases/ICBM152NLin2009).

**Supplementary Figure 3.**
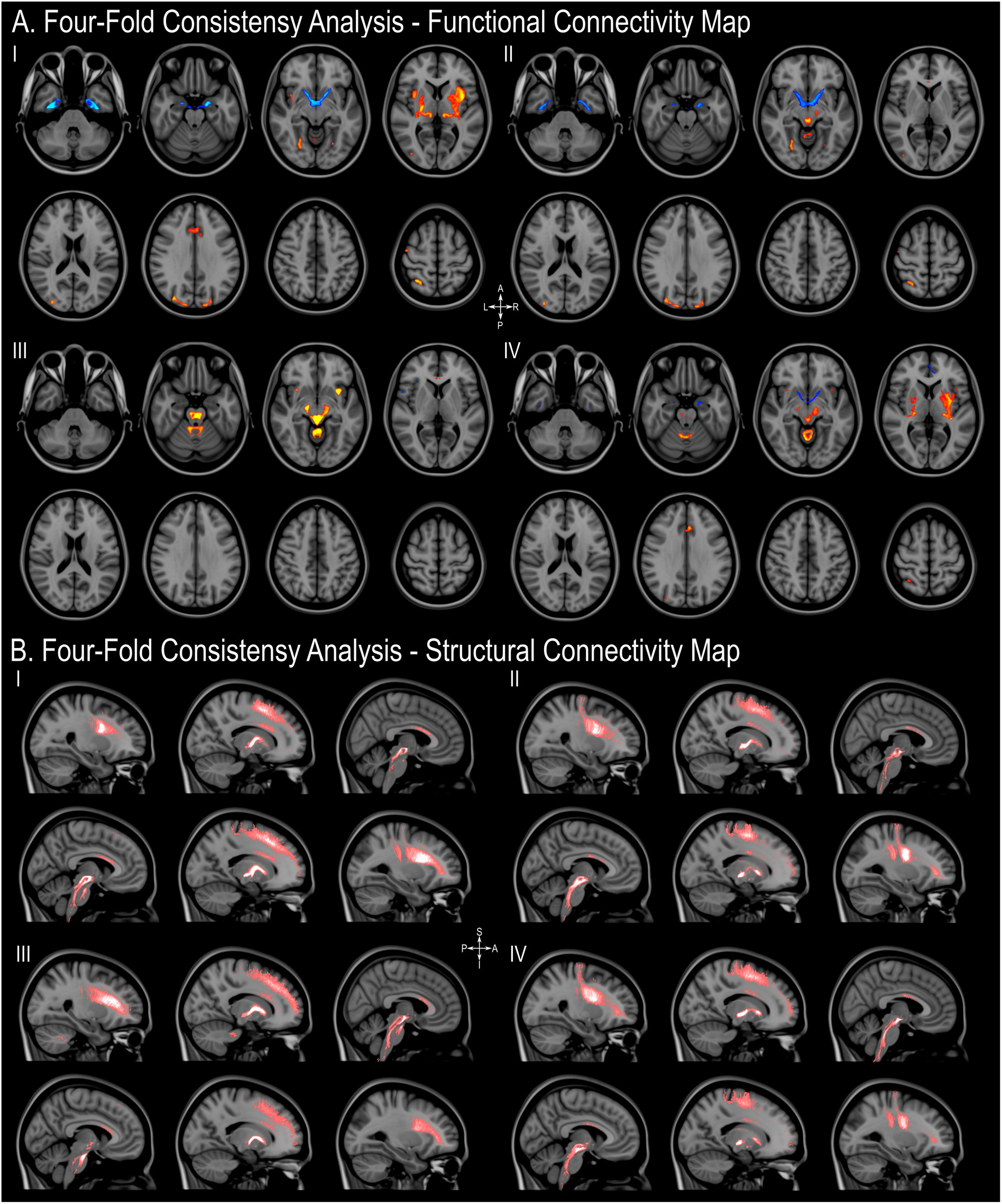
Four fold consistency analysis. To determine the consistency of the results of both structural and functional connectivity mapping, the data was divided in four random groups of even size and the analyses repeated four times leaving out one group each time. While statistical power is reduced due to the smaller sample size in each sub-analysis, the outcomes are highly similar highlighting the same regions and tracts demonstrating that the results reported are consistent across patients. Axial plane (A) and Sagittal plane (B). The results are illustrated in the MNI152 brain (https://www.bic.mni.mcgill.ca/ServicesAtlases/ICBM152NLin2009).

